# Clarity and precision in legal language: Linguistic-discursive analysis of Organic Law 3/2021 on the regulation of euthanasia in Spain

**DOI:** 10.1101/2023.05.18.23290101

**Authors:** A. Hernando-Garreta, C. Centeno, I Olza

## Abstract

Spain has recently regulated the practice of euthanasia through the Organic Law 3/2021. The social relevance of this law clear, as it directly affects fundamental rights enshrined in the Spanish Constitution. The aim of this article is to examine the clarity and precision of the discourse with which the legislator refers to the context of end-of-life care. A linguistic-discursive analysis of the Organic Law 3/2021 is carried out using a mixed methodological approach. The data are categorized and quantified in order to detect potential designative and interpretative problems. We find argumentative inconsistencies in the preamble (n=2) and abundant problematic lexical-semantic choices (n=151) such as vagueness, ambiguity, euphemisms, improper epistemic expressions, evaluative adjectives, verbs of appreciation. Although a legal text must be sufficiently abstract given its generalizable nature, the euthanasia law presents a series of phenomena that affect its interpretation and are potentially negative for its procedural concreteness and effective execution.

## 1. Introduction: Organic Law 3/2021 in Spain

In 1994, the practice of euthanasia^1^ began to be debated in the Spanish Congress and, since then, almost twenty legislative initiatives have emerged over the years. This process recently culminated in the approval of Organic Law 3/2021 (on March 24, 2021) regarding the regulation of euthanasia; it entered into force in June 2021. The context in which this law was drafted and executed is noteworthy in several ways.

(a) On the one hand, the final phase in the process of approving this law coincided with the outbreak of the COVID-19 pandemic, which forced a wide-scale halt of activities and directly impacted the performance of professionals in all areas. It brought about singular –and obvious– complications for workers in the healthcare sector. High rates of death comingled with a feeling of exhaustion on the part of healthcare personnel generated mixed reactions in society and among healthcare professionals themselves when faced with the approval of a euthanasia law that entails specific training for workers in said field (Altisent et al., 2021; Bertolín-Guillén, 2021).

(b) On the other hand, not only was the external context relevant for the drafting and execution of the law, in addition, the period of time after which the regulation itself came into force (three months) was extraordinary since an organic law^2^ of this nature usually takes longer. The *vacatio legis* was scarce in comparison with the training that professionals involved in this matter need, which the law itself proposes (Arruego, 2021). Especially of note, it granted a reduced space of time for the formation of related commissions and the elaboration of the best practices manual that the legislative text proposes.^3^

The elaboration of the law, its implementation and especially its entry into force have generated a complex debate in Spain on a variety of ethical, social and, of course, legal issues. Regular criticism of basic aspects related to the law in terms of its linguistic formulation stands out. Said criticism is found in texts that generally defend the need for the law and its suitability on other levels (Bertolín-Guillén, 2021); it has mostly come from legal specialists generically pointing to the use of euphemisms or ambiguous terms, stressing, above all, the problems that these linguistic phenomena may occasion when applying the law (Martínez López-Muñiz, 2021).

## 2. Objective and structure

Within the framework described, this study aims to present a linguistic-discursive analysis of the text of the law to shed light on its expository clarity and argumentative foundation. This analysis is part of a broader project (author, in progress) to study current public discourse (political, media and institutional) on palliative care (PC) in Spain. It began with analysis of Organic Law 3/2021 since references to PC there strongly impact how the discipline is represented in the public sphere. In a first careful reading of the text, we notice, in line with criticism from jurists mentioned in § 1, what we define *a priori* as inconsistencies and ambiguities in the effective formulation of the law. The present study aims to systematically develop an analysis in this direction, offering structured linguistic-discursive evidence on the expository and argumentative clarity of the law, pointing, above all, to possible interpretive problems that may derive, on the one hand, from the lexical choices of the text and, on the other, from the law’s own argumentative progression. For this, we rely on the premise that all interpretation and application of the law directly depends on its linguistic formulation, as proposed in Article 3 of the Civil Code:

> 1. Regulations will be interpreted *according to the plain meaning contained in their words*, in relation with the context, historical and legislative background, and social reality of the time in which they are to be applied, fundamentally attending to their spirit and purpose” (emphasis added).^4^

Likewise, we start from the hypothesis that, as often happens with legislative texts (Sainz Moreno & Silva Ochoa, 1989), it is possible that Organic Law 3/2021 contains problematic linguistic-discursive choices based on a lack of clarity and precision in the designation of referents, and on deficiencies and weaknesses in its argumentative framework.

After presenting the methodology (§ 3), analysis herein is structured in two large blocks. First, we briefly approach the argumentative body of the preamble (§ 4.1). In what follows, we develop a qualitative critical analysis of passages in the text whose interpretation is subject to ambiguity and vagueness based on certain lexical choices, and to a variable degree of subjectivity, designative non-specificity and/or opacity (§ 4.2). As a result of this study, a series of conclusions are obtained showing that the application of the law may be compromised by deficiencies in its linguistic formulation (§ 5).

## 3. Methodology: Critical discourse analysis and qualitative approaches

This study will be carried out with the hermeneutical approach found in critical discourse analysis (CDA) (Meyer, Wodak, et al., 2003; Pardo Abril, 2013) in order to determine the quality of the text and the possible difficulties social agents may have when interpreting the law regulating euthanasia. It is therefore based on a critical perspective of discourse analysis, which, with sufficient epistemological distance, is capable of understanding and categorizing the text and its interpretation, as well as of detecting possible problematic aspects in its structure and configuration (Gómez García, 2017: 188).

In the framework of the CDA, we will use, fundamentally, qualitative analysis tools, divided into two main facets.

(1) On the one hand, analysis of the argumentative structure will be limited to the preamble, which positions the law socially and legally and establishes the conceptual premises that support the subsequent articles where specific regulatory protocols related to the practice of euthanasia are discussed. This analysis of the preamble of the law is based on the argumentation model defined in van Dijk (1993).

(2) On the other hand, an exhaustive qualitative analysis of the preamble itself and of the rest of the text of the law (articles 1 to 19) will be carried out in order to detect lexical-discursive choices whose interpretation is not unequivocal, or univocal enough. Later, these formulations will be ordered, categorized and quantified in a mixed approach (quantitative-qualitative) that will help identify the passages of the legislative text that give rise to interpretive problems.

## 4. Analysis and results

### 4.1. The argumentation found in the preamble

The preamble to a law is neither mandatory nor regulatory; it therefore allows legislators to flexibly explain the reasons or arguments that found the law and its contents. Despite controversy related to its normative validity, it is traditionally understood as a fundamental source for interpretation of the law. This is so because, as pointed out, the Civil Code establishes that every law must be understood not only in terms of its plain language, but also in the context in which it was issued, and it is precisely in the preamble where legislators present the motivation behind the normative text, as well as its meaning and purpose.

Here, we briefly analyze the argumentative structure of the preamble to the Spanish law regulating euthanasia. Broadly speaking, we find that the preamble follows the structure of legal argumentation formulated in van Dijk (1993), which is presented below and then applied to the arguments contained in the preamble.

This argumentative model allows us not only to situate the conditions under which the text was written (argumentation, justification, framework, circumstances, starting point, facts, legitimacy, reinforcement and conclusion), but also to balance the weight of the arguments and the space dedicated to each of them. The main argumentative problems worth highlighting associated with this preamble are as follows.

(a) The authoritative argument that it cites corresponds to jurisprudence from the European Court of Human Rights, which occupies a greater space than one might, *a priori*, expect since it is used fundamentally to legitimize it even though it lacks a direct thematic relationship with the law (legitimation argument).

(b) The first reason (initial argumentative germ) for which the regulation arises (*responds to demand*) is uncritically assumed and lacks justification since no empirical data is presented that truly demonstrates the need for the norm.

(c) Along these same lines, the fundamental reasons for regulating euthanasia are presented as a series of circumstances that each individual perceives (they are, therefore, not objective: e.g., *enduring unbearable suffering*). Analogous examples from other neighboring countries are also used, yet their similarity to the Spanish case is insufficiently illustrated.

In short, the argumentation on which the is preamble to the law is based is inconsistent in several places, especially considering the absence of objective evidence to justify the so-called “sustained demand in today’s society” on which it is based. This is also clear given the insufficiency of the arguments based on external analogy (e.g., Court of Human Rights, other countries in the European setting) that seek to legitimize it.

### 4.2. Problematic lexical-discursive choices

#### 4.2.1. The quality of legal language

Legal language, and the legal lexicon in particular, have been the object of linguistic study on numerous occasions and the topic of extensive debate. We must first ask for whom laws are written—for the society that must understand their content and abide by then, or for jurists who must interpret them in the first and last instance? If the answer is society, the related texts must be clear and simple such that they are accessible to whomever may receive them; however, if they are addressed to specialists on the matter in order for them to optimally perform their work, inevitably, the language must be specific and technicalities will abound.

Actually, being at the same time addressed to specialists and citizens, legal language should keep a complex balance between clarity and specificity, what allows for an effective and non-problematic application of regulations. Therefore, this language not only has a specific style, but also must adhere to basic stylistic principles such as brevity, simplicity and precision. To the extent that it meets these characteristics, we can assess the quality of the law. However, examples abound of laws that directly contravene these three assumptions. In terms of brevity, we usually find complex syntax that makes reading the law difficult and takes away from simplicity. Regarding precision, ambiguities or vagaries frequently appear that obscure the text and distance it from precision.^5^ In light of these problems, several measures under the scope of Plain Language Movements have been taken to help correct certain linguistic flaws so that citizens can understand what laws say. For instance, Duarte & Martínez (1995) present a small compendium of all the literature or regulations related to drafting laws for their correct elaboration and consequent application, as well as proposals to follow in each one of them. This division was carried by country. Examples of publications regarding legislative drafting include the Austrian guidelines, *Legistische Richlinien*, which stand out for being very thorough, and the German ones, *Handbuch der Rechtsförmlichkeit*, considered the most emblematic. In the Spanish case, perhaps the best known one is the *Acuerdo del Consejo de Ministros* from October 18, 1991, in which guidelines regarding the form and structure of bills were approved.^6^

Despite the fact that, as noted, there has been much effort to improve the quality of drafting laws, the presence of certain linguistic-discursive aspects that make textual interpretation problematic seems inevitable (Marchese, 2012; Rodríguez-Toubes Muñiz, 2017). This is also the case for the Spanish organic law 3/2021, where these problems seem to be more intense at the lexical level, with relevant interpretative and discursive effects that affect clarity in the law.

In what follows, we present a taxonomy of the most common lexical-discursive problems found in the qualitative analysis of the law. Each section (4.2.2-4.2.6) will define, exemplify and critically comment relevant instances of the categories identified, which are not exclusive –i.e. several problems may apply to the same passages and linguistic expressions. In section 4.2.7, a summary of results is offered.

#### 4.2.2. Ambiguity

Ambiguity is opposed to the maxims of precision and clarity and, together with vagueness, constitutes one of the two fundamental problems of legal language, since both contribute to the non-specificity of language. There are three types of ambiguity: semantic, syntactic and pragmatic. Here we are not as interested in deepening the typology of ambiguity as in the specific examples thereof found in the law regulating euthanasia. However, fundamentally, we identify issues of semantic ambiguity, which is, in general, the most productive and least avoidable of the three. By definition, we speak of ambiguity when different –and often contradictory– interpretations of the same word or group of words can be admitted, thus generating confusion and lack of univocal readings.

Let us analyze some examples (emphasis added in all excerpts):

> The legalization and regulation of euthanasia *are based on the compatibility* of certain essential principles that are the foundation of individual rights, and that are thus included in the Spanish Constitution. They are, on the one hand, the fundamental rights to life and physical and moral integrity, and, on the other, constitutionally protected goods like dignity, freedom or autonomy of will.
>
> Making these rights and constitutional principles *compatible is necessary and possible*, for which legislation respectful of all of them is required. (Preamble, page 1)

First, it is stated that the legalization and regulation of euthanasia are based on the compatibility of principles that are the basis of rights and goods and that it is necessary and possible to make those principles and rights compatible. If these principles are the basis of rights, there would be no need to make them compatible, and thus the legislation as proposed would not be necessary. In addition, it seems contradictory to say that it would be necessary and possible to reconcile them if legalization and regulation of the law are based on compatibility. This interpretation abandons the protected goods of dignity, freedom and autonomy of freedom, which, however, later take precedence over the right to life. This is not a legal problem, but rather a problem in how it was drafted, and can cause confusion.

On the other hand, the following statement also seems ambiguous:

> To that end, this law regulates and decriminalizes euthanasia *in certain clearly defined cases* that are subject to sufficient guarantees that safeguard absolute freedom of decision, ruling out external pressure of any kind. (Preamble, page 2)

It is ambiguous to speak of certain cases if the law contemplates legality only in the case of serious, chronic and disabling disease or serious and incurable illness as defined in Article 3. The phrase *certain cases* implies a broader and more difficult to interpret series of possibilities given the lack of specificity. This question could have been easily solved by referring to the fundamental requirement of serious, chronic and disabling disease or serious and incurable illness and then expanding the conditions to which they are subject. Increasing the casuistry in which euthanasia is acceptable seems intentional; this also fits in with the guaranteeing nature of the law, which safeguards the different particularities in society.

On the other hand, at the end of the preamble, recapitulating the need to regulate the practice of euthanasia and all that it implies, we find two other ambiguities that generate gaps and may incite certain debate, namely *mitigated by other means* and *a fully capable and free person*.

> It is understood as an action that directly and intentionally causes the death of a person through a single and immediate cause-effect relationship, at the informed, express and repeated request of said person; it is carried out in a context of suffering due to an incurable illness or disease that the person experiences as intolerable and that could not be *mitigated by other means*. (Preamble, page 3)

What other means are referred to here? If strictly applied, this seems to imply that a patient with an incurable illness or disease will not be able, for example, to request euthanasia if they have not previously received palliative care and have not yet exhausted all possibilities of relief. However, we know that this is not the case because, Article 5, in *Requirements for the provision of assistance in dying* states that *the patient must have all available information in writing regarding their medical process, the different alternatives and possibilities of action, including access to comprehensive palliative care*… However, having said information in no way obliges them to receive these services. Data collected by research in palliative medicine reveals reduced access to this type of care for a variety of reasons.^7^

On the other hand, and further on, after considering constitutional rights and goods in the context of euthanasia, the patient is required to be a *fully capable and free person* in order to precedence over the right to life:

> When a *fully capable and free person* faces a vital situation that, in his opinion, violates his dignity, intimacy and integrity, as defined by the context of euthanasia described above, the good of life can wane in favor of other goods and rights with which it must be weighed, since there is no constitutional duty to impose or protect life at all costs and against the will of the bearer of the right to life. (Preamble, page 3)

Based on the context, this statement seems to exclude all people who have some sort of intellectual disability or a disease that directly affects said abilities. However, Article 6, referring to the *Requirements for requesting the provision of assistance in dying*, contemplates different solutions so that they can access it in case of incapacity. Another problem that stems from this ambiguity is the use of the adjective *free*. It seems evident that, when a person is ill and experiences intolerable suffering, (s)he cannot be free since (s)he is under the pressure of pain and (s)he may be surround by other circumstances that do not allow him/her to freely decide. This would be the case of those who feel they are a burden to their family, do not have financial resources to pay treatment, or are in an anxious-depressive state, etc.

Another case of ambiguity is found in the definition of *Informed Consent*:

> Informed consent: the patient’s free, voluntary and conscious agreement, expressed in full use of their faculties after receiving the *appropriate information*, so that, at their request, one of the actions described in letter g) takes place. (Article 3, a), page 4)

On what basis is this information adequate? Clearly this includes reference to the information that their doctor should provide about prognosis, treatments, etc., but what may be adequate for the doctor could be insufficient for the family, for example. Specifying this in more detail may help serve as a guide for the medical community and thus help avoid possible communication problems or those related to the doctor-patient relationship.

Another definition that could generate controversy is that of *Situation of de facto incapacity*, Article 3, h), precisely because, as in the previous example, the lack of specification can generate ambiguity:

> “Situation of de facto incapacity:” a situation in which the patient lacks sufficient understanding and will to autonomously, fully and effectively govern his or herself, regardless of whether *support measures* are or have been adopted for the exercise of their juridical capacity. (Article 3, h), page 5)

What type of support is necessary in this case? From the context, this likely refers to legal measures, however, some might think it refers to psychological support measures for the patient prior to their incapacitation. A person outside the world of law would not necessarily know which documents or actions this statement refers to. The syntactic use of the passive voice (measures have been adopted) is also complex, although common in the drafting of laws. Who is directly responsible for this issue? The text also fails to provide clues regarding the citizen who must interpret the law, which generates certain confusion or difficulty when applying the law.

Regarding the wording of Article 4 on the *Right to request the provision of assistance in dying*, we repeatedly find another series of ambiguities that generate gaps or, rather, favor non-specificity towards increasing the euthanasia context and guarantee greater access. These expressions or phrases include: *means of support and resources will be guaranteed; accessibility measures; reasonable modifications; pertinent measures to provide access*. The nouns *means, resources, measures* and *modifications* are all polysemic and open up a wide field of possibilities. In addition, in the last two examples, the adjectives that specify them, *reasonable and pertinent*, imply subjectivity due to their evaluative nature so they do not help to clarify the terms.

Articles 5 and 6, which are dedicated to the requirements for receiving the provision and making the request respectively, contain other examples of ambiguity and both specifically refer to *means*.

> Having voluntarily made two requests in writing, *or by other means* that leave a record, and that are not the result of any external pressure, with at least fifteen calendar days between each request. (Article 5, c), page 5)

The only alternative to a request in writing would be an oral one, which we understand as a request in the form of a voice recording, video or similar. However, it could also just be a simple conversation. Only requiring that it *leaves a record* means anyone could prepare a written text that would substantiate a conversation about the person’s will and thus provide the required evidence, but they could have changed the information based on specific interests at play. The same can be found in Article 6:

> In the event that it is impossible for you to date and sign the document due to your personal situation or health condition, you may use *other means* to record it, or *another person* of legal age and fully capable may date and sign it in your presence. (Article 6, 1), page 6)^8^

There is a clear lack of specificity about *the means* for *leaving proof* because using another means, as expressed, might give proof of a request, but it would not necessarily include a signature. The same would happen with *another person*. The use of indefinite determiners like *another* generates precisely what its name indicates, indefiniteness. Since this is a law that seeks to guarantee and establish a new right, it is reasonable to think that its use of words is deliberate and contemplates all possible particularities. However, using this resource is sometimes a double-edged sword for jurists, who, due to their lack of specification, may face interpretive problems.

#### 4.2.3. Vagueness

Vagueness supposes, along with ambiguity, one of the biggest problems when interpreting the law, as mentioned in the previous section. A term is vague when it does not have a specific determination, but rather is broad or general and is also free in its choice or application. The imprecision or indeterminacy that characterize vagueness generates an unspecific context of interpretation that is sometimes conflictive.

When talking about vagueness, in general, a distinction is made between two types, namely intensional vagueness and extensional vagueness. For the former, indeterminacy does not allow the question to be known exactly. An example of this is the world *relevant*. For extensional vagueness, however, there is some gradation. An example in this sense can be found in Article 11, 1), referring to the *fulfilment of the provision of assistance in dying*.

> Once positive resolution has been reached, healthcare professionals must fulfil the provision of assistance in dying with the utmost care and professionalism, applying the corresponding protocols, which will also contain *criteria regarding how and when to carry out the request*. (Article 11, 1), page 8)^9^

The expression of containing *criteria regarding and when* is vague because it does not specify if certain and orderly guidelines must be followed, nor in relation to time are periods, hours or minutes spoken of. Here interpretation and calibration of the margins in this regard are up to those responsible for execution of the protocols.

When dealing with the phenomenon of vagueness, it is also common to differentiate between gradual and combinatorial vagueness when carried out from the point of view of legal interpretation. Another possible classification of vagueness refers to the dimensions of the term as part of analysis from the philosophy of language (Ferrari (2020) and Keefe (2000) respectively).

From the philological point of view, the vagueness in the *Euthanasia Regulation Law* is closely linked to the subjectivity that underlies the terms and that is so recurrent in the text.

#### 4.2.4. Subjectivity

Legal-administrative texts are mainly characterized by objectivity since clarity and precision are sought, and, for this, a denotative lexicon must be used, that is, an objective one. However, in the law that concerns us, the presence of subjectivity is striking, especially that which is manifested through evaluative adjectives –whose interpretation relies on each subject’s perspective– and the verbal semantics. This subjectivity also results in certain fundamental terms being vague. Thus, we find, for example, *unbearable suffering* a fundamental requirement for the practice of euthanasia to take place. This *suffering* is subject to diverse perspectives depending on the case. What is unbearable for one patient will not be the same or in the same quantity for another, although the context may be the same or similar. That one of the fundamental requirements is problematic reveals a deliberate use of language and discloses the quality of the law.

The examples listed in the table below help illustrate the subjectivity in the text. Nominal syntagma (NS) are collected in the left-hand column and verbal syntagma (VS) in the right-hand column. Isolated words organized by grammatical categories are not presented because, in this way, it is easier to contextualize the terms and analyze their subjectivity load, as well as their positive or negative connotations.

Taking into account NS, it first seems evident that a negative connotation prevails. However, there are more nouns with a neutral or positive connotation, like *environment, decisions, improvement*, when faced with repeated suffering. This is because the adjectives that specify them, except for appreciable, are all negative. Thus, the reader’s interpretation constitutes a problem that should be resolved regardless of whether it is a subjective question or not. An apparent exception to these examples is found in the NS phrase *appreciable improvement*, but if we contextualize it, we find that it speaks of the non-existence of appreciable improvement, so semantically it also refers to an outwardly negative context.

The term *unfavorable environment* is also a vague and subjective concept. One’s environment may be determined by very different circumstances and are not specified. Thus, for example, a patient’s family context, economic situation, personal baggage and current situation, etc. all come to mind. The key, however, lies in what it means to be unfavorable. We understand unfavorable for situations that present an adverse and detrimental scenario in all cases where there are objective signs of said adversity. However, in the context of illness, what might seem irrelevant *a priori*, could be an unbearable burden for the patient. Measuring or contemplating these cases is a problem for the legislator given the impossibility of collecting the entire range of possibilities. Therefore, given the nature of the subject, these vague terms are intentionally used without specification and indirectly favor the expansion of the euthanasia context.

Verbal syntagma, for its part, contains degrees of subjectivity that are complemented, for the most part, with an evaluative adjective implying an attributive or predicative function.

The inverted pyramid represents the subjective load that verbs themselves contain. With *to consider* and *to experience,* the individual develops the action, making it more subjective; on the other hand, with *to emerge, to be* and *to exist*, the individual is not an agent, but rather simply expresses an attitude towards it and there is less subjectivity in the syntagma. If we look at the examples presented, we see that the verb *to consider*, the most subjective of all, is by far the most productive. The adjectives with which the verbal nuclei are combined are ordered from negative to positive valencies.^11^ Undoubtedly, all of them are evaluative and that they all involve some subjectivity. For this reason, establishing clear and objective delimitations in the requirements for applying for euthanasia is a complex task. The arguments in the preamble entail less objectivity when evaluative language is present.

#### 4.2.5. Euphemisms

Sometimes, the difficulty to interpret legislative language is based on the presence of euphemisms, which contribute to an overall strategy of mitigation of negative effects in communication. Replacing negative and taboo terms with counterparts that have a positive connotation attenuates the impact of a possibly threatening message.

In the law analyzed here, there are a series of euphemisms to replace the expression *deliberate act to end life*, which necessarily appears in the text in order to define euthanasia. The euphemistic terms used are varied and include *assistance*, *act of medical aid*, *provision*, or *healthcare action*, which have a rather positive connotation. We briefly collect the results obtained when analyzing euphemisms in the law.

The left-hand column presents the euphemisms or euphemistic expressions used and their raw frequency. The right-hand column presents the use of non-euphemistic terms or direct references to death. The overall numerical difference is very high. We found 108 euphemistic phrases compared to 4 direct references to death, revealing a clear intention to get rid of *death* as a term, considering it taboo. The two most frequent substitutions are *euthanasia* (21) and *provision of assistance in dying* (58); the rest of the expression directly related to one of the two. There are two constructions with reference to euthanasia (*euthanasia-related conduct* and *euthanasia-related act*), while the rest are a simplification or version of *provision of assistance in dying*. Perhaps more surprising is the fact that the law was not named the *Regulation of the provision of assistance in dying* instead of the *Regulation of euthanasia*. Perhaps this decision was carried out following the example of other countries that previously decriminalized the practice.

In any case, it seems that these uses are linked to a consistent will to evade in order to hide an unpleasant idea, namely death. In this sense, some authors believe that, when employing euphemisms, it is not so much a question of attenuating the message, but of expressing what is considered politically correct. On the other hand, euphemisms are sometimes resorted to as a manipulation mechanism through linguistic obscurity with ambiguous or vague words or constructions. As we have shown, this also occurred in the drafting of this law, where, on occasions, delimiting certain issues becomes particularly complex or gives rise to a variety of interpretations. This communicative strategy contravenes the maxims of clarity, accuracy and conciseness, but it is diluted by other bureaucratic transfers and, for many, goes unnoticed (Sánchez García: 2018, 31-89).

Finally, let us examine one last complex example. In the preamble, euthanasia is defined as a *deliberate act of ending a person’s life*, but later it is said to be an act of medical aid. This is not possible since a doctor seeks to eliminate suffering rather than the person, which is what has generated controversy. A euphemism is acceptable as long as it does not contravene the case. In other examples, however, we can clearly see how a new reality is constituted through euphemistic language, such as the fact that deliberate death is not only not intended, but is now a provision, an aid and, ultimately, a right.

#### 4.2.6. False synonymy

Finally, another linguistic issue that often generates confusion when interpreting the law refers to proposed synonymies between terms that are not, in fact, synonymous. In the case of LO 3/2021, it becomes especially problematic given that it occurs in the definition of terms that are a *sine qua non* condition for accessing euthanasia. They include *grave, chronic and disabling illness* and *grave and incurable disease*. Both *illness* and *disease* are partial synonyms that give rise to confusion between the two and, therefore, a broad definition of each is offered. However, the problem persists.

Regarding definitions, there are a series of basic rules and guidelines to follow when drafting legislation that in the text under discussion have not been complied with and may cause confusion.^12^ According to Salvador Coderch (1989: 166-167), it is advisable to “*define only if necessary*. A legislator’s definitions tend to specify cases of vagueness, settle doubts in cases of ambiguity, restrict or expand usual meanings, particularize the reference to a certain set of things, etc.”. We start from the fact that the law that concerns us here surely intended to avoid vagueness, ambiguity and to specify, however, vague or ambiguous terms were used in the definition that do not resolve the issue. For example, a *grave, chronic and disabling illness* is defined as a *situation that refers to limitations*… In order to clarify them and resolve the situation, adjectives are used, but as advanced in the subjectivity section, far from specifying, it broadens the question to the perspective of the person who experiences and assesses it. The definition thus fails to meet the aim and becomes invalid.^13^

Another basic guideline to follow includes not defining the same expression differently when using it several times (Salvador Coderch, 1989: 169-170). In the development of this specific guideline, it is necessary to use different terms to designate different concepts. This is clearly neglected since *serious illness* and *serious disease* are defined. It would have been more appropriate to use different adjectives to help the reader more clearly differentiate since the terms are non-specific and synonymous.

To these two problematic examples we can add other drafting issues. The two concepts in question sometimes appear in one order (Preamble, Article 3 and First Final Provision) and sometimes in another (Articles 5 and 12). In addition, there is sometimes mention of *physical or mental suffering* (for example: *with the consequent physical or mental suffering* in Preamble I) and *physical and psychological suffering* at other times (for example: *causing physical or psychological suffering* in Preamble II). In context, we clearly understand that this is about the same thing, but the synonymy between mental and psychological was unnecessary and only contributes to the problem. Let us recall that these are the fundamental prerequisite for requesting euthanasia, which is why the drafters should have taken special care when using them and should have attempted to avoid causing confusion.

#### 4.2.7. Summary of results

Below we include a synoptic table of the mixed analysis carried out in this study. The main lexical-discursive problems detected through qualitative methods are summarized and then broadly quantified. The main argumentative inconsistencies analyzed in section 4.1 are also included in the table. The full analysis of the law is provided as supplementary material in: https://bit.ly/euthanasiaSpain.

## 5. Discussion

The linguistic difficulties that can arise in drafting a given law are evident and, despite the efforts of professionals to avoid them, adapting as much as possible to the regulations governing legislative language, a variety of problems arise that make it difficult to interpret the text. As seen in this linguistic-discursive analysis, *Organic Law 3/2021 (March 24) on the regulation of euthanasia* presents problematic issues related to linguistic drafting and, therefore, interpretation.

Firstly, the present study has confirmed that the preamble to the law conforms to the pattern of legal argumentation found in van Dijk, but it has also revealed certain elements that leave room for debate. The arguments are not always backed by evidence and the text does not meet standards of clarity and precision.

Secondly, the law as a whole presents more than forty examples of linguistic issues that lead to non-specificity and, therefore, give rise to interpretative issues. Its ambiguity, vagueness and subjectivity very often leave gaps that, together with euphemisms and false synonyms, obscure the message and raise the possibility that the law intends to broaden the euthanasia context. On the other hand, given the nature of the subject matter, which incites diverse reactions, a deliberate use of language to mitigate is evident. All of these linguistic issues denote a hasty drafting of the law and generate interpretive problems that directly affect the correct application of the law.

We can conclude, therefore, that this legislative text presents a series of discourse-related inconsistencies that are possibly the result of both the rapid context in which the law was elaborated, and the typical nature of legal language. To regulate the practice of euthanasia, surrounding countries have faced a similar problem due the use of non-specific language. For instance, in the recent case of Portugal (2021), the law was only approved after having to reformulate the text.^14^ This fact once more highlights the need for linguistic-discursive analysis of legislative texts for their corresponding approval, interpretation and application.

This research did not receive any specific grant from funding agencies in the public, commercial, or not-for-profit sectors.

## Data Availability

All relevant data are within the manuscript and its Supporting Information files.

https://bit.ly/euthanasiaSpain.

## Notes

1. The mixed parliamentary group that year, with deputy Pilar Rahola as spokesperson, promoted a non-legal proposal for the creation of a parliamentary paper with the aim of studying “legal recourse for the social demand that has emerged around euthanasia.”

2. Note here the third final provision of the law, where the ordinary nature of certain articles and provisions is specified: “This law has the character of an organic law with the exception of articles 12, 16.1, 17 and 18, of the first additional provisions, second, third, fourth, fifth, sixth and seventh, and the sole transitory provision, which have the character of ordinary law”.

3. For this reason, it was, in fact, necessary that Article 17, referring to the Guarantee and Evaluation Commissions, enter into force –exceptionally– the day after the publication of the law in the BOE. It would be impossible to practice euthanasia without these commissions and without a best practices manual to ensure compliance.

4. Original Spanish text: “Las normas se interpretarán según el sentido propio de sus palabras, en relación con el contexto, los antecedentes históricos y legislativos, y la realidad social del tiempo en que han de ser aplicadas, atendiendo fundamentalmente al espíritu y finalidad de aquellas”.

5. Other typical characteristics are, for example, a technical lexicon, a tendency to nominalization, overuse of passive constructions or the use of identifying determiners.

6. This example is repealed by a subsequent Agreement: BOE 180/2005, of July 29, 2005, with reference BOE-A-05-13020. Also of note, the Report of the Commission for the modernization of juridical language presented in 2011 to the Council of Ministers and accessible at the General Access Point of the Administration of Justice.

7. See official data for Spain in the European Atlas of Palliative Care. Spain has 0.6 specialized services per 100,000 inhabitants, while the European average is 0.8. The optimal ratio, according to the standards of the European Association for Palliative Care, is 2. (Arias-Casais et al., 2019. *EAPC Atlas of Palliative Care in Europe 2019*).

8. Original Spanish text: En el caso de que por su situación personal o condición de salud no le fuera posible fechar y firmar el documento, podrá hacer uso de *otros medios* que le permitan dejar constancia, o bien *otra persona* mayor de edad y plenamente capaz podrá fecharlo y firmarlo en su presencia.

9. Original Spanish text: Una vez recibida la resolución positiva, la realización de la prestación de ayuda para morir debe hacerse con el máximo cuidado y profesionalidad por parte de los profesionales sanitarios, con aplicación de los protocolos correspondientes, que contendrán, además, criterios en cuanto a la forma y tiempo de realización de la prestación.

10. There is no parallel equivalence between the verbal actions and the adjectives presented in the table.

11. Salvador Coderch (1989) defines a set of norms and guidelines to follow for elaborating definitions in legislation. We will exclusively focus on the two guidelines that are most relevant here.

12. It seems widely accepted that, when drafting a law, some intentionally leave it as such, postponing the matter until a judge rules, if the need to specify arises.

13. See, for example: “*Portugal’s top court rejects bill to legalise euthanasia*. […] The decision came after the country’s recently re-elected President Marcelo Rebelo de Sousa, a conservative, asked the court to evaluate the legislation on the grounds that *it appeared to contain “excessively undefined concepts”.* Article published in *Reuters* (March 15, 2021), https://www.reuters.com/article/us-portugal-rights-euthanasia-idUSKBN2B72JP.

---

**Figure 1.**
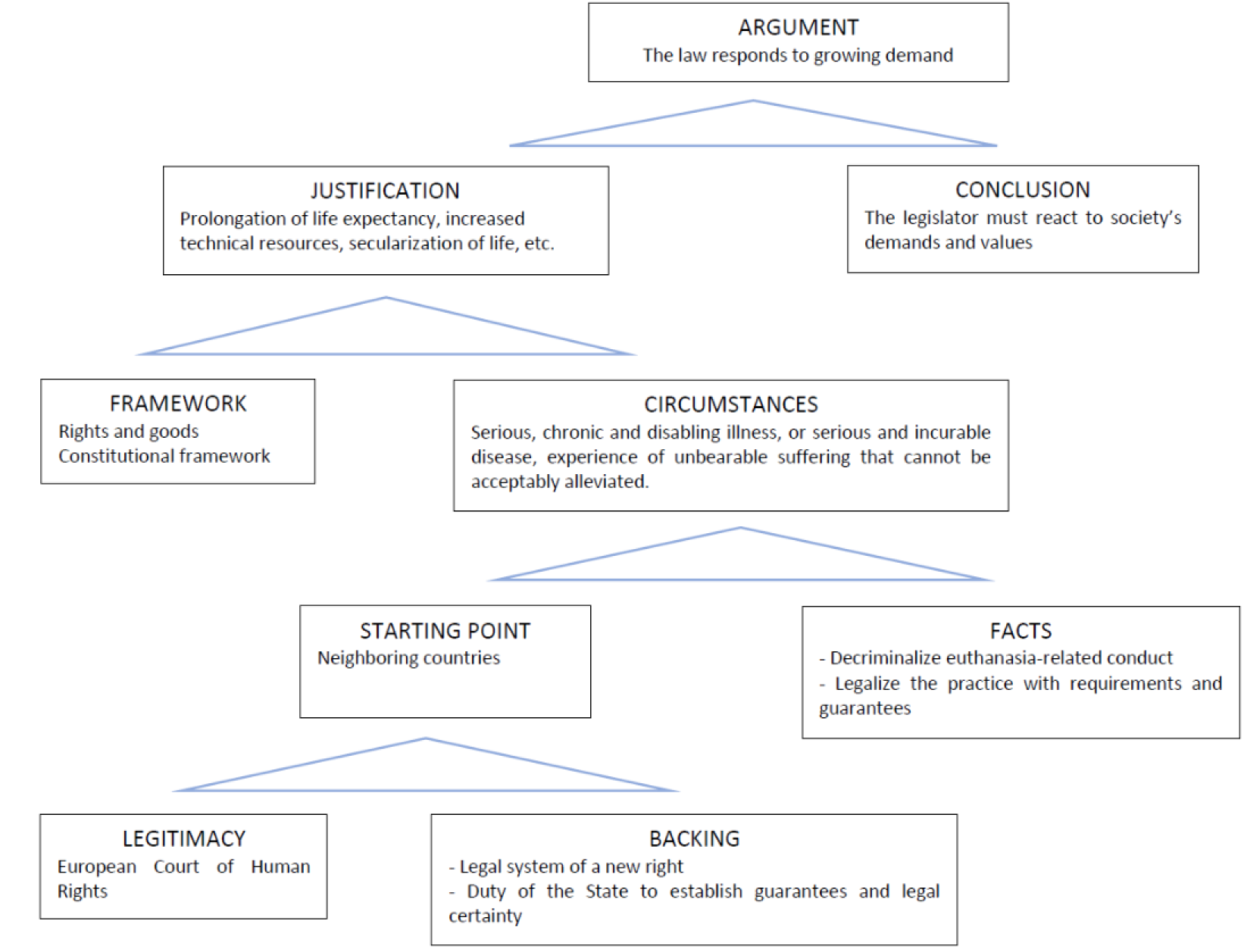
Argumentative structure of the preamble to the law

**Figure 2.**
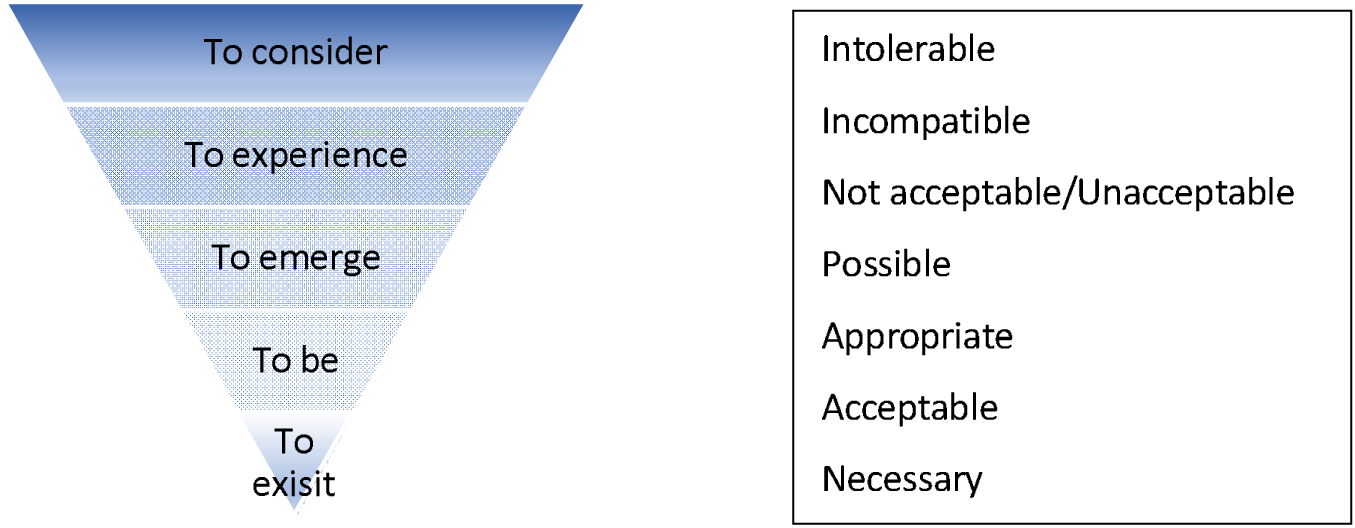
Subjective load and evaluative valency of verbs and adjectives

**Table 1.** Adjectives and verbs based on subjective interpretations

| **Adjectives (NS)** | **Verbs (VS)** |
| --- | --- |
| *Unbearable suffering* | *It is necessary and possible* |
| *Unfavorable environment* | *Considered acceptable* |
| *Hasty decisions* | *Considered unacceptable* |
| *Appreciable improvement* | *Considered incompatible* |
| *Constant and intolerable suffering* | *Experienced as unacceptable* |
|  | *There is a high probability* |
|  | *The doctor believes that* |
|  | *Considered intolerable* |
|  | *It is incompatible* |
|  | *Considered appropriate* |
|  | *Considered without prospects* |

**Table 2.** Euphemisms and non-euphemistic expressions

| EUPHEMISMS OR EUPHEMISTIC EXPRESSIONS |  | NON-EUPHEMISTIC EXPRESSIONS |  |
| --- | --- | --- | --- |
| *Assistance in death* | 1 | *Deliberate act to end life* | 1 |
| *Assistance necessary to die* | 2 | *Deliberate end to a patient's life* | 1 |
| *Assistance in dying* | 7 | *To end life* | 1 |
| *Provision of assistance in dying* | 58 | *Action that causes a person's death* | 1 |
| *Provide assistance in dying* | 1 |  |  |
| *Assistance* | 2 |  |  |
| *Provision* | 8 |  |  |
| *Act of medical aid* | 1 |  |  |
| *Healthcare action* | 1 |  |  |
| *Euthanasia-related conduct* | 5 |  |  |
| *Euthanasia-related act* | 1 |  |  |
| *Euthanasia* | 21 |  |  |

**Table 3.** Summary of results.

| Phenomenon | (#) | Category | Linguistic effect | Example |
| --- | --- | --- | --- | --- |
| Mitigation | 108 | Euphemisms | Opacity and imprecision | *Act of medical assistance, healthcare action, aid in dying, aid for dying, aid for death, service, aid, euthanasia-related conduct, euthanasia-related action, euthanasia* |
| Non-specificity | (>40) | Vagueness | Imprecision | *Incurable disease or illness that a person experiences as unacceptable and that has not been alleviated by **other means**.* |
|  |  | Ambiguity | Imprecision | *The legalization and regulation of euthanasia **are based on the compatibility** of certain essential principles... including, on the one hand, the fundamental rights to life and to physical and moral integrity, and on the other, constitutionally protected goods such as dignity, freedom and autonomy of will. It is **necessary and possible** to make these constitutional rights and principles compatible, and it requires legislation that respects all of them.* |
|  |  | Subjectivity | Imprecision | Evaluative adjectives: ***unbearable, unfavourable, hasty, appreciable, constant, intolerable*** |
|  |  |  | Imprecision | Verbs of appreciation: ***to consider, to experience, to emerge, to be (+adj) and to exist (+adj).*** |
|  |  |  | Imprecision | Epistemic expressions: ***in their view*** |
| Argumentative inconsistency | 4/6 | Absent or unspecific argumentative evidence | Argumentative weakness | *And it is precisely the legislator's obligation to respond to society's demands and **values**, preserving and respecting rights and adapting the rules that order and organize our coexistence.* |
|  |  | Questionable wording | Argumentative weakness | *The European Court of Human Rights' ruling from May 4, 2013 (Gross v. Switzerland) is relevant in that it considers it **unacceptable that a country that has decriminalized euthanasia-related conduct does not develop and enact a specific legal regime**, specifying the modes of practice of said euthanasia-related conduct.* |

## Notes

### Competing Interest Statement

The authors have declared no competing interest.

### Funding Statement

The authors received no specific funding for this work

## References

Studies

Altisent, R., Nabal, M., Muñoz, P., Ferrer, S., Delgado-Marroquín, M. T. & Alonso A. Eutanasia: ¿es esta la ley que necesitamos? Atención Primaria; 2021. 53, 102057. http://dx.doi.org/10.1016/j.aprim.2021.102057

Arias-Casais, N., Garralda, E., Rhee, J. Y. et al. EAPC atlas of palliative care in Europe. Romania; 2019. 122, 0–6. https://dadun.unav.edu/handle/10171/56787

Arruego, G. Las coordenadas de la Ley Orgánica de regulación de la eutanasia. Revista Española de Derecho Constitucional; 2021.122, 85–118. https://doi.org/10.18042/cepc/redc.122.03

Author (in progress).

Bertolín-Guillén, J. M. Eutanasia, suicidio asistido y psiquiatría. Revista de la Asociación Española de Neuropsiquiatría; 2021. 41(140), 51–67. https://dx.doi.org/10.4321/s0211-57352021000200003.

Duarte, C., & Martínez, A. El lenguaje jurídico. Buenos Aires: A-Z; 1995.

Dijk, T. A. van. Principles of critical discourse analysis, Discourse and Society; 1993. 4(2), 249–283.

Ferrari Yaunner, M. El lenguaje del Derecho, retos y posibilidades para la interpretación jurídica. Revista Derechos En Acción; 2020. 17(17). https://doi.org/10.24215/25251678e476

Gómez García, J. A. La argumentación jurídica: teoría y práctica. Madrid: Dykinson; 2017.

Keefe, R. Theories of vagueness. Cambridge: Cambridge University Press; 2000.

Marchese, M. C. El discurso legal-normativo como praxis social. Análisis crítico de las leyes sobre vivienda para habitantes de la Ciudad de Buenos Aires en situación de pobreza. Revista de Llengua i Dret. Journal of Language and Law; 2012. 57, 43–70.

Martínez López-Muñiz, J. L. El deber de proteger la vida, y especialmente de los más debilitados, frente a un inexistente derecho a quitarse la vida por sí o por otros. Revista Española de Derecho Constitucional; 2021. 122, 47–83. https://doi.org/10.18042/cepc/redc.122.02

Meyer, M., Wodak, R., Eguibar, B. & Fernández Aúz, T. Métodos de análisis crítico del discurso. Barcelona: Gedisa; 2003.

Pardo Abril, N. Cómo hacer análisis crítico del discurso. Una perspectiva latinoamericana. 2a. edición. Universidad Nacional de Colombia: Instituto de Estudios en Comunicación y Cultura (IECO); 2013.

Rodríguez-Toubes Muñiz, J. La imprecisión del lenguaje legislativo, expuesta en el artículo 18 LRJSP. Cuadernos Electrónicos de Filosofía Del Derecho; 2017. 36, 11. https://doi.org/10.7203/CEFD.36.10447

Sainz Moreno, F., & Silva Ochoa, J. C. da. La calidad de las leyes. Vitoria-Gasteiz: Parlamento Vasco; 1989.

Salvador Coderch, P. Definiciones y remisiones. In Sainz Moreno, F., & Silva Ochoa, J. C. da. (1989): La calidad de las leyes. Vitoria-Gasteiz: Parlamento Vasco; 1989. 157–182.

Sánchez García, F. J. Eufemismos del discurso político: las claves lingüísticas del arte del disimulo. Madrid: Visor Libros; 2018.

Legislative and Institutional texts

Código Civil español. Real Decreto de 24 de julio de 1889 por el que se publica el Código Civil. Gaceta de Madrid. Núm. 206, de 25 de julio de 1889. https://www.boe.es/eli/es/rd/1889/07/24/(1)/con

Ley Orgánica 3/2021, de 24 de marzo, de regulación de la eutanasia. Boletín Oficial del Estado, núm. 72, de 25 de marzo de 2021. https://www.boe.es/diario_boe/txt.php?id=BOE-A-2021-4628

Proposición no de Ley sobre la creación de una ponencia que estudie dar curso legal a la demanda social generada en torno a la eutanasia (núm. exp. 162/000093). Pleno del Congreso de los Diputados (24 abril 1994). https://app.congreso.es/AudiovisualCongreso/escucharAudio?legislatura=Legislatura_V&carpeta=Iniciativas&nombreFich=05_000400_19940426_01_162000093.mp3&tipo=S

Resolución de 28 de julio de 2005, de la Subsecretaría, por la que se da publicidad al Acuerdo del Consejo de Ministros, de 22 de julio de 2005, por el que se aprueban las Directrices de técnica normativa. Boletín Oficial del Estado, núm. 180 de 29 de julio de 2005. https://www.boe.es/eli/es/res/2005/07/28/(1)/con

